# An Evaluation of Methods for Monitoring Annual Quality Measures by Month to Predict Year-End Values

**DOI:** 10.1101/2021.01.18.21250069

**Authors:** Todd R Johnson, Krishna Kumar Kookal, R Joseph Applegate, Suhasini Bangar, Alfa Yansane, Joel M White, Ryan Brandon, Kristen Simmons, Joanna Mullins, Ana Neumann, Muhammad F Walji, Elsbeth Kalenderian

## Abstract

**Background:** An increasing number of healthcare quality measures are designed for annual reporting. These measures require an entire year of data to accurately report the percentage of patients who met the measure. Annual measures give providers latitude to prioritize clinical workload and patient needs; however, they do not provide a direct means to monitor performance throughout the reporting year. Although there are many possible methods for measuring annual measures at finer-grained timescales, our applied work showed that the most obvious methods could give a misleading and inaccurate view of progress throughout the year. Neither the definitions of the annual measures, nor the research literature, provided any guidance on the best methods for interim monitoring of annual measures.

**Objective:** Our objective was to evaluate four different methods for monitoring annually reported quality measures monthly to best predict year-end performance throughout the reporting year.

**Methods:** We developed four methods for monitoring annual measures by month: 1) Monthly Proportion: The proportion of patients with one or more encounters in the month who still needed to meet the measure at their first encounter of the month and met the measure by the end of the month; (2) Monthly Lookback Proportion: The proportion of patients seen in the month who met the measure by the end of the month, regardless of whether it was met in that month or previously in the reporting year; (3) Rolling 12 Month: The annual measure reported as if each month was the twelfth month of a twelve-month reporting period; and (4) YTD (Year-to-Date) Cumulative: The proportion of patients with one or more visits from the start of the reporting year through the month who satisfy the measure. We applied each method to two annual dental quality measures using data from two reporting years, and four different dental sites. We used mean squared error (MSE) to evaluate year-end predictive performance.

**Results:** Method 3 (Rolling 12 Month) had the lowest MSE in 11 out of 16 cases (2 measures X 2 years X 4 sites) and lowest total MSE (262.39) across all 16 cases. In 5 of the 16 cases, YTD Cumulative had the lowest MSE.

**Conclusions:** The Rolling 12 Month method was best for predicting the year-end value across both measures and all four sites.

## Introduction

Initiatives to assess and improve outpatient healthcare quality increasingly rely on annually reported measures that require data from an entire reporting year. For example, the diabetic oral exam measure [1] reports the percentage of diabetic patients who had an oral exam within the reporting year. Annually reported measures are common in outpatient care because they reflect the clinical reality that patient needs and clinical workloads may delay screenings, or that some measures, such as blood pressure control, may only be addressed over time and several visits. Although annual measures provide a long-term view of healthcare quality, providers need a means to identify measures for quality improvement initiatives, and a means to assess those initiatives at finer-grained time scales. Unfortunately, none of the annual measures that we have seen to date provide guidance on how to monitor interim progress throughout the year, nor could we find any guidance in the literature. In addition, our experience with over 60 different annually reported measures suggests that the best method will likely depend on both the nature of the measure and the goals of those who need to track the measure.

For instance, suppose that we create a monthly report in which the numerator is the number of patients who received a diabetic oral exam at any time from the start of the reporting year to the end of the month and the denominator is the number of diabetic patients who visited at least once in that same time. The monthly percentage reported for the last month of the reporting year will necessarily match the annual measure (since the annual measure considers diabetic patients who visited at any time during the year). However, due to clinical workload or clinical concerns about proper spacing of screenings, patients who visit early in a new reporting year may not meet the measure after his or her first, or even first few, visits. When graphed by month, we have found that this method often shows a large performance decrement at the start of a new reporting year, followed by steady improvement. The apparent decrement is a result of delaying screenings until later visits. The apparent improvement arises from the fact that patients tend to visit more than once in the reporting year, which means that more and more patients meet the measure over time. With this monthly measure, neither the decrement nor the improvement necessarily reflects a decrease or increase in the screening rate.

Now consider reporting the measure each month by considering in the denominator only those diabetic patients who had a visit in the month and still required screening. Let the numerator be the number of those patients who received the screening in that month. This monthly view of the annual measure more closely tracks the actual monthly screening rate. If the goal is to assess an intervention designed to improve the screening rate, this might be a good measure of monthly performance. However, this method seems unlikely to give a good indication of the final year-end value, because each month looks only at patients who visit that month and still require a screening. It does not consider patients who visited in previous months and either had or did not have the screening. As a result, the reported monthly screening rates are unlikely to align with the annual screening rate.

The objective of this research was to empirically evaluate four different methods for monitoring annual, once-a-year measures monthly, to best predict year-end performance. Once-a-year measures are those that require each eligible patient to meet a condition one time in the reporting year. In this paper, we focus on two measures: an annual diabetic oral exam[1,2] and an annual caries risk assessment.[3] Other similar measures in the overall health arena include annual falls screening for elderly patients[4] and annual flu vaccination.[5]

## Methods

We developed four methods for monitoring annually reported measures by month. These methods were based on existing methods that we have seen used in our own and other quality improvement initiatives. The methods, along with short descriptors used in the remainder of the article, are: 1) Monthly Proportion: The proportion of patients with one or more encounters in the month who still needed to meet the measure at their first encounter of the month and met the measure by the end of the month; (2) Monthly Lookback Proportion: The proportion of patients seen in the month who met the measure by the end of the month, regardless of whether it was met in that month or previously in the reporting year; (3) Rolling 12 Month: The annual measure reported as if each month was the twelfth month of a twelve-month reporting period; and (4) YTD (Year-To-Date) Cumulative: The proportion of patients with one or more visits from the start of the reporting year through the month who satisfy the measure.

We applied each monitoring method to the specifications for the annual diabetic oral exam[2] and the annual caries risk assessment[3] measures separately to data from three US dental schools and one large dental accountable care organization (Sites A to D in the results) for 2016 and 2017. Since each site in this study used the same Electronic Dental Record (EDR) system, we developed and validated SQL scripts for each method at one site. Validation was done in two steps. First, a dentist at the site where the scripts were developed validated the data by randomly picking patients from each measure and reviewing the patient charts in the EDR. We then distributed the scripts to the other three sites to execute them on their EDR. The scripts took the reporting year as input and queried the underlying EDR database to produce a list of patients who met the measure criteria for the numerator and denominator based on the definition of each measure and each method. The output of the SQL script included an identifier for the institution, the patient’s Medical Record Number (MRN), a deidentified patient identifier for analysis and visualization purposes, the measure name, the method name, the reporting year and month and the patient’s demographic information. We then conducted a second validation of the results of each method from each site by cross checking for mathematical consistency within and across methods. For example, Rolling 12 Month and YTD Cumulative must both produce the final annual measure for month 12, hence their values should match. Likewise, the values for the first month of each annual reporting period should be equal for all methods other than Rolling 12 Month. We used an iterative process to review, revise, and rerun the scripts to ensure that each method was correctly applied to each measure at each site.

The output of the SQL scripts were combined into one SQL database table. We used Tableau Desktop Version 2018.3.16 for visualization and initial analysis. To measure how well each method predicted year-end performance, we used the mean squared error (MSE) of each monthly result with respect to the final year-end value. The performance of methods for predicting year-end values may be affected by large changes in the annual measure from one period to the next. To determine whether a site’s performance on a measure statistically differed btween 2016 and 2017, we used R Version 3.6.3[6] to conduct a two-sided z-test for difference of proportions (α = .05).

## Results

Figure 1 shows the results of each method for the two measures across all sites and years. Each subplot shows the monthly trend for each of the 4 methods for a single site, measure, and year. The gray horizontal line in each subplot shows the final year-end value of the measure. The subplots illustrate some similarities and differences across methods, as well as across the two measures. We discuss these in the next section.

**Figure 1:**
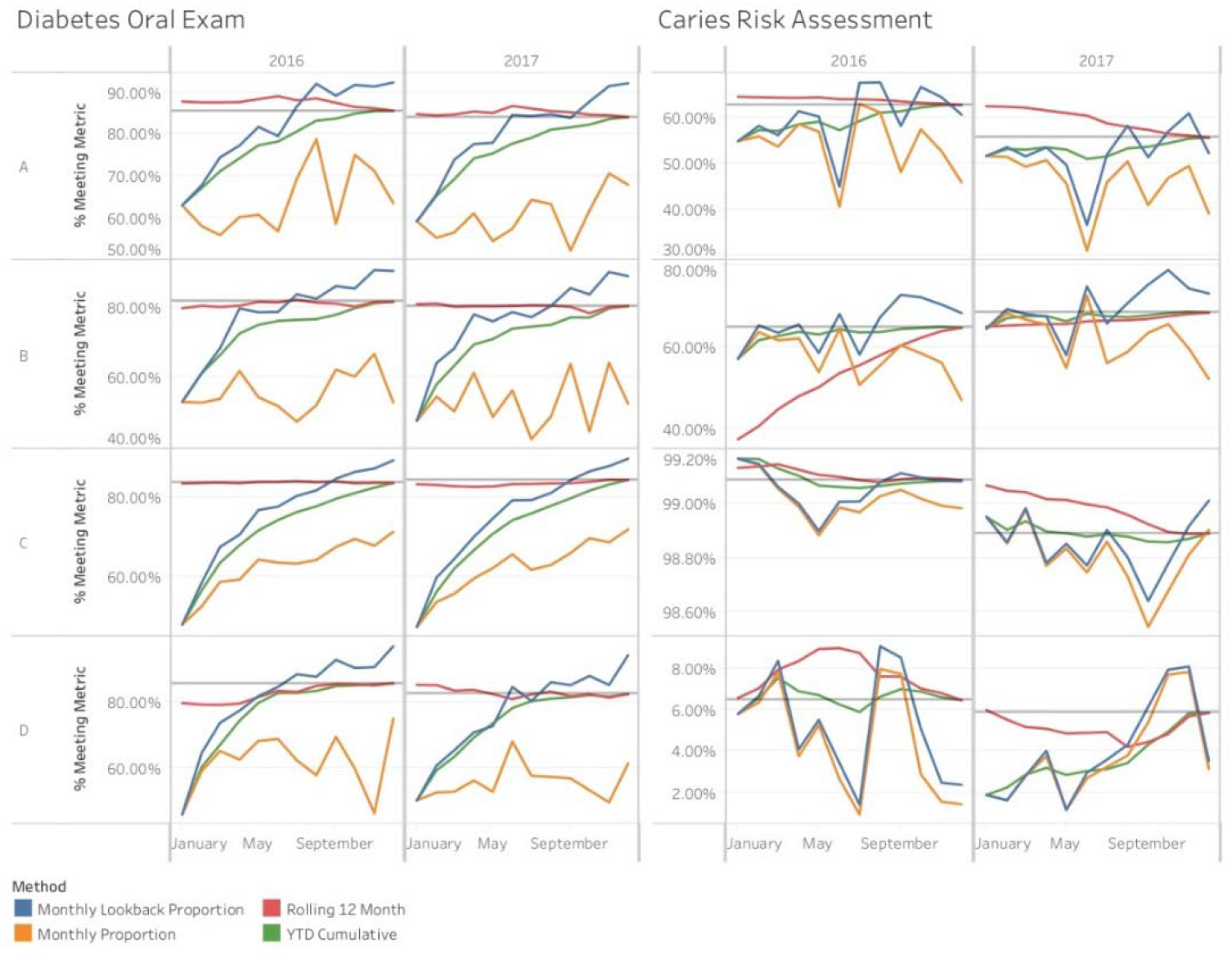
Graphical results of the four methods for all sites, measures, and years. To better depict differences across methods, Y axes are independent for each site and measure.

Table 1 shows the MSE of each method by site. Rolling 12 Month had the lowest MSE in 11 out of 16 cases (2 measures X 2 years X 4 sites), while in the other 5 cases YTD Cumulative had the lowest MSE. The table also shows the grand total of the MSEs for each method across all sites, years, and measures. Here, Rolling 12 Month showed the best performance with a total MSE of 262.39, followed by Monthly Lookback Proportion with a total MSE of 1,482.32. YTD Cumulative’s total MSE of 1,542.03 was close to that of Rolling 12 Month, whereas Monthly Proportion fell far behind all other methods with a MSE of 5,402.06.

**Table 1:** Mean squared error of monthly predictions to the final year end value for each measure, method, year and site. The lowest MSE in each row is displayed in italics.

|  |  |  | Monthly<br>Lookback<br>Proportion | Monthly<br>Proportion | Rolling 12<br>Month | YTD<br>Cumulative |
| --- | --- | --- | --- | --- | --- | --- |
| Measure | Year | Site |  |  |  |  |
| Diabetes Oral<br>Exam | 2016 | A | 104.0187 | 509.5843 | 4.7548 | 112.4926 |
|  |  | B | 135.1035 | 694.3542 | 0.8696 | 140.0362 |
|  |  | C | 211.2270 | 499.9471 | 0.0530 | 254.7321 |
|  |  | D | 201.1187 | 627.7408 | 15.2391 | 223.0994 |
|  | 2017 | A | 106.8147 | 594.7106 | 1.5970 | 121.1737 |
|  |  | B | 143.0702 | 798.9109 | 0.4267 | 182.1742 |
|  |  | C | 236.5896 | 555.6842 | 1.2605 | 289.9967 |
|  |  | D | 180.5714 | 715.0110 | 1.8110 | 178.7387 |
| Caries Risk<br>Assessment | 2016 | A | 45.2171 | 113.5457 | 1.5754 | 16.8871 |
|  |  | B | 26.9591 | 75.8140 | 206.8081 | 6.3476 |
|  |  | C | 0.0055 | 0.0096 | 0.0009 | 0.0014 |
|  |  | D | 7.5509 | 10.1402 | 2.2277 | 0.2259 |
|  | 2017 | A | 43.3420 | 127.7016 | 19.8734 | 7.5376 |
|  |  | B | 32.5193 | 70.1713 | 5.0792 | 2.1100 |
|  |  | C | 0.0115 | 0.0206 | 0.0116 | 0.0007 |
|  |  | D | 8.2022 | 8.7177 | 0.8067 | 6.4763 |
| Grand Total |  |  | 1,482.32 | 5,402.06 | 262.39 | 1,542.03 |

Table 2 shows the year end values for each measure by site. Only the Caries Risk Assessment measure for sites A, B, and C differed statistically across the two years. Note that these cells in Table 1 were 3 of the 5 cells where YTD Cumulative outperformed Rolling 12 Month.

**Table 2:**
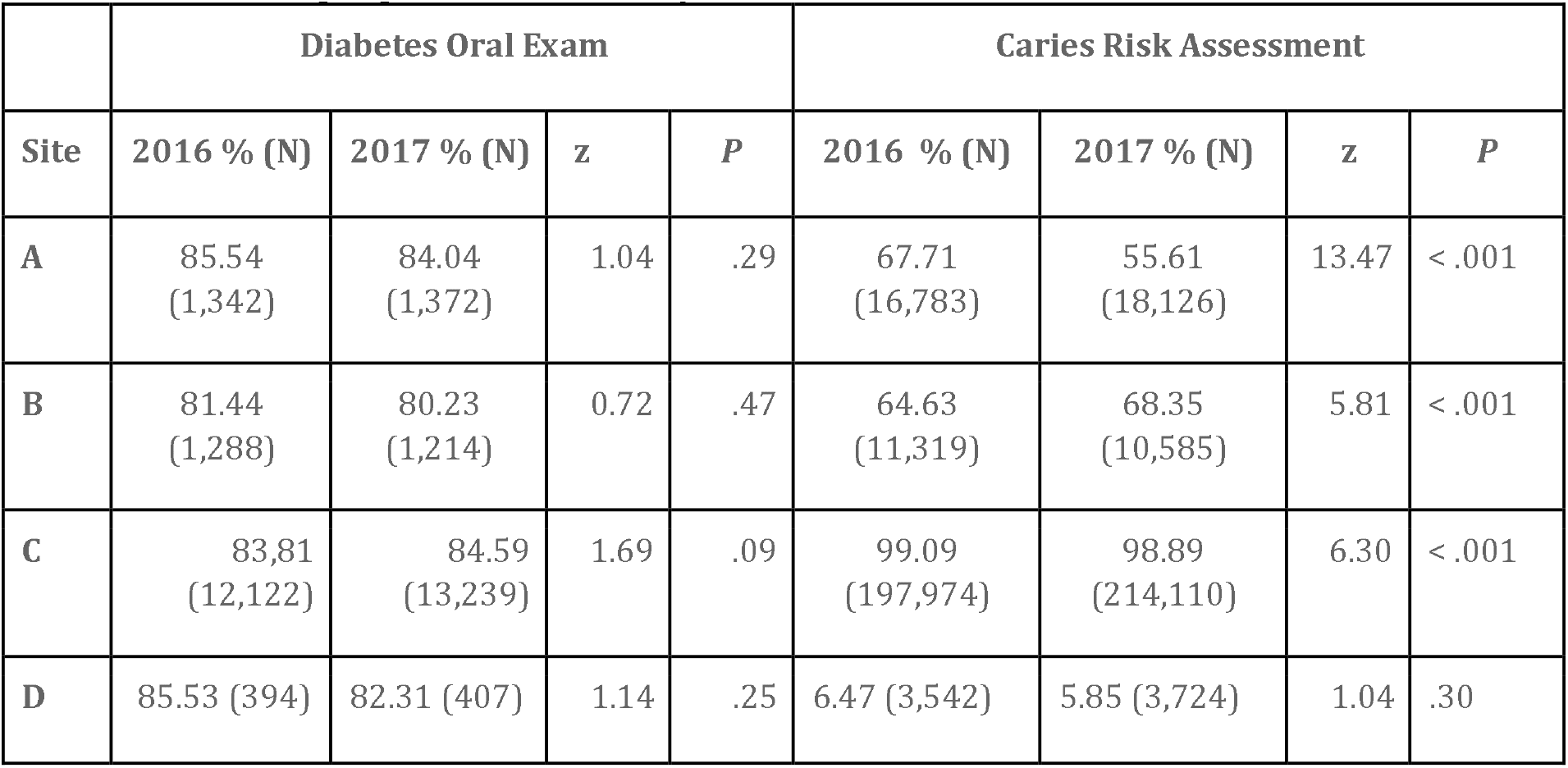
Year end results for each measure, site and year and results of a Z-test for differences in proportion between years.

## Discussion

Across the 16 cases, Rolling 12 Month was the most consistent method for closely predicting year-end results, with the lowest MSE in 11 of the 16 cases and the lowest total MSE. In the 5 cases where YTD Cumulative had a lower MSE than Rolling 12 Month, Rolling 12 Month was second best in 4 cases. In 3 of those 4 cases, YTD Cumulative was just slightly better than Rolling 12 Month. However, in one case, Caries Risk Assessment, 2016 Site B (See Table 1), Rolling 12 Month performed far worse than all other methods with an MSE of 206.81 versus the next highest at 75.81. To explore this outlier, we modified the SQL scripts for Site B to examine its 2015 data for Caries Risk Assessment. The year-end results for 2015 and 2016, respectively were 34.51% and 64.63%, a statistically significant difference at the .95 level. Since Rolling 12 Month looks back 11 months, it is not surprising that this sudden jump in performance led to a high MSE for Rolling 12 Month in 2016. Thus one limitation of Rolling 12 Month is that any sudden major shift in performance on a once-a-year measure is likely to result in a higher MSE than the other methods. Another limitation is that it requires complete data from the previous reporting year.

Rolling 12 Month’s limitation for predicting year-end values in the face of large shifts in measure performance suggests that modifying the method to more heavily weight recent values may improve its performance. One common approach is to use an exponentially weighted moving average (EWMA) chart[5] that gives more weight to recent values. However, EWMA averages values from the current and several previous months, whereas Rolling 12 Month does not average the past 12 months, but instead reports a monthly value as if that month were the end of a 12 month reporting period. Any application of EWMA to Rolling 12 Month would lead to a new monitoring method. One obvious approach is to apply EWMA to the monthly values produced by Rolling 12 Month. Although this might result in a method with improved performance, it introduces a number of complexities that may not be worth any possible improvement in performance. First, EWMA charts may be more difficult for stakeholders to interpret since each monthly value will depend on the primary method for monitoring the annual measure by month and the average of the current and past months. Second, EWMA requires an additional parameter, *λ*, to control the weighting of each value. For instance, if *λ*= 0.2, the weight of the most recent value is given 0.2 and the weights of the previous values are 0.16, 0.128, and so forth.[7] The performance of any approach using EWMA will thus depend on the method used to compute the monthly values and the value of *λ*.

Monthly Lookback Proportion and Monthly Proportion consistently performed much worse than Rolling 12 Month and YTD Cumulative. Although Monthly Lookback Proportion was a distant second best in terms of total MSE, it was never the best method for any of the 16 cases. Monthly Proportion fared the worst in terms of MSE, making it the worst method for predicting year-end results. However, Monthly Proportion is the most accurate method, by definition, for monitoring the performance on a measure in a given month, because it measures the percentage of patients who visited in that month while still missing the measure, who then met the measure by the end of the month. As a result, while Monthly Proportion does not predict year-end performance well, it may work well for detecting sudden shifts in performance throughout the year. Another limitation of Monthly Lookback Proportion and Monthly Proportion is that the value for the final month of the year does not typically reflect the final year-end value of the 12 month reporting period, because both methods are based only on patients seen in a given month. As shown in Figure 1, both methods are often far from the final year end value (indicated by the horizontal lines) for all 16 cases.

In contrast to Monthly Lookback Proportion and Monthly Proportion, Rolling 12 month and YTD Cumulative are guaranteed (by definition) to end on the final yearly value at the end of Month 12 of the reporting year. However, in many cases, the monthly values for YTD Cumulative tend to be low at the start of the reporting year, then gradually climb to the year-end value. Figure 1 shows this pattern in 13 of 16 cases. This is due to the once-a-year requirement of the measure and the fact that at the beginning of a new reporting year, all eligible patients no longer meet the measure. For various clinical, workflow, or workload related reasons, not all patients who have visited in the first month will receive the screening required to meet the measure. However, each month that a patient returns to a practice there is a new chance for the patient to meet the measure. Once met, a patient is in the numerator for all remaining months. Consider the preventive care oral exam measure and assume that in a given month the probability of doing the preventive care oral exam for a patient seen in that month is *r*. The probability, *p*, of a patient being screened after *n* visits is:

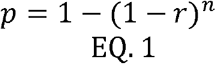

Figure 2 shows the probability of meeting a measure after 1 to 10 visits for several different values of *r*. This mirrors the pattern seen for YTD Cumulative in 13 of the 16 cases. Monthly Lookback Proportion shows a similar pattern in 11 of the 16 cases, since it too reports only on patients who visited in the month. As a result, Monthly Lookback Proportion and YTD Cumulative often give a misleadingly low estimate of actual performance early in the year (see Figures 1 and 2) which may lead the viewer to incorrectly assume that performance has suddenly dropped.

**Figure 2:**
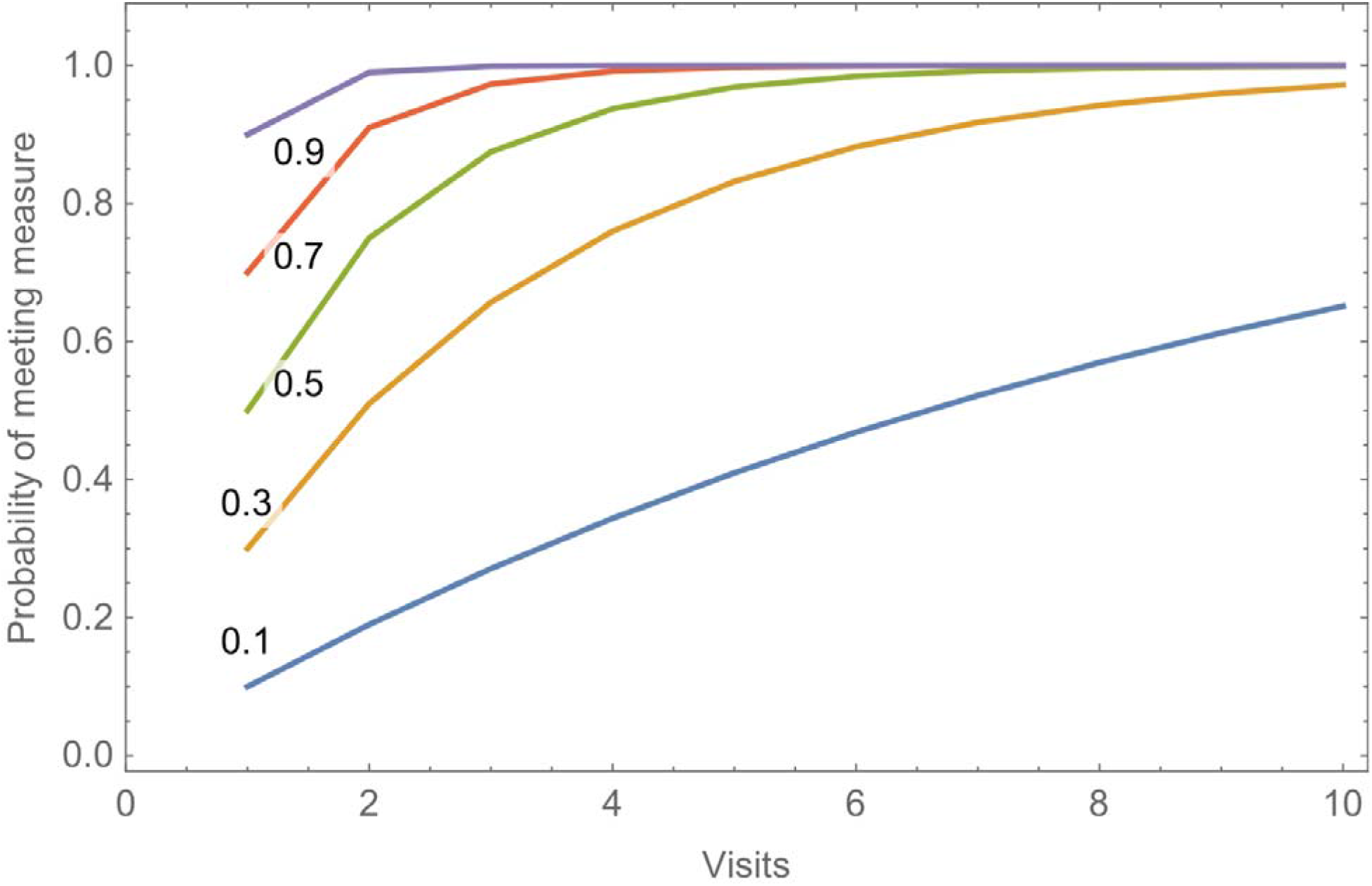
Probability of meeting a once a year measure as a function of the number of visits (1-10), if the probability of being screened in a visit is r, where r ranges from 0.1 to 0.9.

One limitation of this study is that it evaluates methods only for once-a-year measures. These types of measures have two unique properties. First, once the patient meets the measure, that patient has met the measure for the entire reporting year. Second, if a patient does not meet the measure at the beginning of a visit, action taken in that visit can guarantee that the patient meets the measure by the visit’s end. Other types of measures lack these properties. For instance, NQF’s (National Quality Forum) measure 0018, Controlling High Blood Pressure,[6] is based on a patient’s blood pressure from his or her most recent visit, hence as the year progresses a patient may fall in and out of the measure. In addition, if a patient’s blood pressure is too high at a visit (assuming it was properly measured), there is no way to improve it at that visit. Improvement might only be detected months or more after an initial visit. We call measures such as this, episodic measures to reflect the notion that a patient can fall in and out of the measure at each episode of care. Such temporal constraints often vary widely across measures. The NQF 2902 measure for postpartum contraceptive care[8] measures the percentage of patients who received appropriate contraception within 60 days of a live birth, but excludes patients who gave birth in the last two months of the reporting period, since those patients may not have the opportunity to receive contraception. These differing temporal constraints may require different methods for accurately predicting year-end values. Indeed, it raises the question if once-a-year-measures are appropriate for such temporally constrained measures.

A second limitation is that this study looks only at methods for predicting the final year-end value, whereas quality improvement activities also require a means to detect changes in performance throughout the reporting period. Change detection may be used to determine where improvement is needed, to assess the success of a quality improvement initiative, or to monitor a process for unexpected changes in performance. Clinical decision support systems may be developed to direct the appropriate and timely patient care that predict success for the annual measure. Additional future work is needed to examine a combination of the method used to monitor an annual measure at a finer time scale together with a specific statistical approach for detecting change. In our operational work we have used X-Charts,[7] year-over-year graphs with Z-tests, and Analysis of Means for Proportions Charts,[9] however, many other change detection approaches are available, such as EWMA and CUSUM (cumulative sum) charts.[7] In addition, the appropriate change detection method may depend on the temporal characteristics of the measure.

Finally, we have not considered how best to display progress on measures to stakeholders throughout the year. Organizations often use reports or live dashboards to convey performance measures throughout the reporting period. These may be used by various stakeholders, including leadership, quality improvement staff, and clinical staff to gauge and adjust performance. In some cases, these measures are reported at subunit and even individual clinician levels with statistical comparisons of units at these lower levels of the organization. Although Rolling 12 Month was best at predicting year-end values, it is not likely the best method for conveying to stakeholders a clear picture of how well the organization or subunits are doing at any point in time. In our operational work, we have found that stakeholders prefer YTD Cumulative, once they understand why that method may suddenly drop at the start of a new reporting period. Detailed user studies are needed in this area so as to develop evidence-based guidelines for conveying measure progress to stakeholders throughout the reporting year and the possible positive impact timely and accurate measure progress may have on patient care

## Conclusions

The increasing emphasis and importance on healthcare quality measures requires clear evidence-based guidelines that stakeholders can use to predict and assess performance on measures throughout and across reporting periods. In this study, we evaluated four methods for monitoring annual once-a-year measures at the monthly level in order to best predict year-end results. The data showed that the Rolling 12 Month method, in which each month is treated as if it was the end of a 12 month reporting period, performed best across the 16 cases (2 measures X 2 years X 4 sites). Additional research is needed to assess the best approach for measures that are not once-a-year and for change detection throughout the reporting period.

## Data Availability

Data will be made available upon reasonable request.

## Acknowledgements

All authors approved the submitted paper and made a substantial contribution to the conception or design of the work, and to the acquisition, analysis or interpretation of the data. Research reported in this article was supported by award R01DE024166 from the National Institute of Dental and Craniofacial Research, National Institutes of Health. The content is solely the responsibility of the authors and does not necessarily represent the official views of the National Institutes of Health. This study was approved by the Committee for the Protection of Human Subjects at each institution. UTHealth acted as the main IRB where the study is approved under protocol number HSC-DB-14-1051: Implementing Dental Quality Measures in Practice. De-identified data are available upon reasonable request.

## Abbreviations

YTD: year-to-date
MSE: mean squared error
EWMA: exponentially weighted moving average
NQF: National Quality Forum
CUSUM: cumulative sum

